# Impact of school closures on the health and well-being of primary school children in Wales UK; a routine data linkage study using the HAPPEN survey (2018-2020)

**DOI:** 10.1101/2021.02.04.21251155

**Authors:** Michaela James, Emily Marchant, Margaret A. Defeyter, Jayne V. Woodside, Sinead Brophy

## Abstract

**Introduction:** In response to the COVID-19 pandemic, school closures were implemented across the United Kingdom. This study aimed to explore the impact of school closures on children’s health by comparing health and wellbeing outcomes collected during school closures (April – June 2020) with data from the same period in 2019 and 2018.

**Methods:** Data were collected online via the ‘HAPPEN At Home’ survey, which captured the typical health behaviours of children aged 8 – 11 years between April - June 2020. These data were compared with data in 2018 and 2019 also collected between April-June, from HAPPEN. Free school meal (FSM) status was used as a proxy for socio-economic deprivation. Analyses were repeated stratifying by FSM.

**Results:** Comparing responses between April – June in 2020 (n=1068), 2019 (n=1150) and 2018 (n=475), there were improvements in physical activity levels, sleep time, happiness and general wellbeing for children during school closures compared to previous years. However, children on FSM ate less fruit and vegetables (21% (95%CI (5.7% to 37%)) and had lower self-assessed school competence compared to 2019. Compared to those not on FSM they also spent less time doing physical activity (13.03% (95%CI: 3.3% to 21.7%) and consumed more takeaways (16.3% (95%CI: 2%-30%)) during school closures.

**Conclusion:** This study suggests that schools play an important role in reducing inequalities in physical health. The physical health (e.g. physical activity and diet) of children eligible for FSM may be impacted by prolonged school closures.

**What is already known on this subject?:** In response to the COVID-19 pandemic, by mid-March 2020, 138 countries had implemented national school closures to reduce the number of social contacts between pupils, therefore interrupting the transmission of COVID-19 as part of pandemic plans. UNESCO warned that the global scale and speed of the educational disruption would be unparalleled. There is an ongoing debate with regard to the effectiveness of school’s closures on transmission rates, but the fact schools are closed for a long period of time could have detrimental impacts on pupil’s physical and mental health.

This study provides evidence of any differences in the health and wellbeing of children prior to and during the COVID-19 enforced lockdown and school closures between March and June 2020. These findings could have a significant impact for the future and support schools to better understand their pupil’s physical, psychological, emotional and social health. It also contributes to a significant literature gap regarding the impact of school closures on school-aged children.

**What this study adds?:** Improvements in physical activity levels, sleep time, happiness and general wellbeing were observed in general for children during school closures compared to previous years. However, children on FSM reported eating less fruit and vegetables and had lower self-assessed school competence compared to 2019. Compared to those not on FSM they also spent less time doing physical activity and consumed more takeaways during school closures. These trends are not evident among children not on FSM. All children reported improvements in wellbeing during lockdown especially on the happiness with family measure.

Overall, findings suggest schools help to reduce inequalities in physical health for socio-economically deprived children. During school closures children from deprived backgrounds are likely to have poorer physical health (e.g. less time spent doing physical activities and poorer diet) and this is not observed in children who are not in receipt of FSM. This research suggests that school closures will result in widening health inequalities and when schools return measures will need to be in place to readdress the widened gap in physical health.

## Introduction

In early March 2020, the World Health Organisation (WHO) declared the coronavirus disease (COVID-19) to be a global pandemic^1,2^. To reduce the risk of person-to-person transmission, a wide range of public health measures were implemented by governments worldwide. These included the closure of educational settings in order to reduce the number of social contacts between pupils^3,4^. By April 2020, the United Nation’s Educational, Scientific and Cultural Organisation (UNESCO) estimated that 138 countries had implemented national school closures, impacting around 80% of children worldwide^4^.

There is an ongoing debate regarding the effectiveness of schools closures on transmission rates^4–6^ but the fact schools were closed for a long period of time could have had detrimental impacts on pupil’s physical and mental health^4,5,7,8^. School closures may have reduced opportunities for physical activity, extracurricular activities, school meals and social interaction ^9–12^. Research shows that when children are out of school (e.g. weekends and holidays) they are less physically active, have longer screen time, irregular sleep patterns, less favourable diets, weight gain and a loss of cardiorespiratory fitness^5,13^. This is noted to be particularly detrimental for those from more deprived backgrounds^4,6,10,12^.

A report by the Royal Society’s Data Evaluation and Learning for Viral Epidemics (DELVE) group highlighted concerns regarding the increased inequalities in children’s physical and mental health as a result of school closures^14^. For example, pre-existing inequalities such as food poverty are likely to be exacerbated through reduced access to free school meals^15^. Thus, there is a real possibility that, in addition to a widening of the educational attainment gap, school closures are likely to result in widening inequalities in children’s physical health, mental wellbeing, and health related behaviours.

This study aims to; 1) compare children’s health and wellbeing during school closures in 2020 with the same period in 2019 and 2018 and, 2) stratify the before and during period of school closures by socio-economic deprivation (as measured by free school meal (FSM) eligibility).

## Methods

### Study Design

The HAPPEN Wales network was established at Swansea University in 2015 following research with headteachers who advocated for collaboration and a joined up approach to prioritising health and wellbeing within the school setting^16^. The network involves children aged 8–11 years completing the HAPPEN Survey, an online self-report questionnaire that was developed and designed with children. The survey captures a range of information on health and wellbeing including nutrition, physical activity, sleep, wellbeing and concentration^17^. Prior to school closures, children completed the survey within the school setting during curriculum time. A data collection and feedback system is achieved by sharing group-level results to schools as a school report tailored to the curriculum. Annual reports are also shared with key stakeholders in health and education.

In light of the COVID-19 pandemic, HAPPEN aimed to understand how school closures were affecting the health and wellbeing of children in Wales. Therefore, the original HAPPEN Survey was adapted to the ‘HAPPEN At Home’ survey to capture changes in health behaviours due to school closures and provide schools the opportunity to gain a better understanding of pupil’s health and wellbeing. This enabled schools to plan for and address any concerns they identified within their ‘HAPPEN At Home’ report during the return to school. The survey was granted ethical approval by Swansea University’s Medical School on 15/04/2020 (Reference: 2017-0033B).

### Participants

Recruitment of participants and data collection was delivered online due to COVID-19 restrictions. Pre-existing HAPPEN schools were emailed initially inviting them to participate in the ‘HAPPEN At Home’ survey. Next the survey was then opened wider and all primary schools in Wales were contacted through a number of methods including direct email, a social media campaign (paid advertisement on Facebook and Twitter) and promotion from key stakeholders (e.g. regional education consortia). Schools were invited to share details of the survey (including study aims and information sheet) amongst parents/guardians so that children could complete the survey at home at a convenient time. Communication between schools and parents/guardians was achieved through existing channels such as text messages, newsletters and social media. This is the same sampling method as the 2019 data however, 2018 data was collected in South Wales as the network was not pan-Wales in 2018.

### Data Collection

Primary data were collected via the ‘HAPPEN At Home’ Survey between April and June 2020. The survey captured the typical health behaviours of children aged 8-11. Items included validated measures of physical activity, sedentary time, diet and dental health^18^, as well as wellbeing, competency and autonomy. Items included in the analyses are presented as supplementary information (S1). The full versions of the ‘HAPPEN At Home’ and original HAPPEN survey can be viewed in the supplementary information (S2 and S3 respectively).

The survey was conducted online and could be completed by children at home or in school (key worker or vulnerable children) via mobile phone, tablet and computer. The process of data coding involved two researchers. The first researcher downloaded the raw data, cleaned the data, checked for duplicates, generated a unique participant ID number and removed identifiable information. This process protects participants’ anonymity by ensuring that the second researcher generating the report and conducting the analysis could not identify individuals. Raw data was coded using STATA (version 16) to produce a dataset for the purpose of analyses.

Free school meal (FSM) status was used as a proxy for deprivation^19^ and was obtained via the Secure Anonymised Information Linkage (SAIL) Databank^20^. To link the data, the demographic data are separated from the responses and sent to a trusted third party, NHS Wales Informatics Service (NWIS) and the response data goes to SAIL using a secure file upload. A unique Anonymous Linking Field (ALF) is assigned to the person-based record before it is joined to clinical data via a system linking field.

### Analysis

Primary analysis looked at whole group mean comparison of all children from 2018 and 2019 (pre-school closures) to 2020 (school closures). Secondary analysis included the subset of children from 2019-2020 stratified by FSM. The 2018 data was used to account for annual trends prior to lockdown.

For this paper, school closure was categorised as the period between 20^th^ March 2020; the date in which the Minister for Education in Wales set for the closure of statutory education provision and 29^th^ June 2020; the date in which schools returned for a phased approach in Wales. The ‘HAPPEN At Home’ survey was launched in 23^rd^ April 2020 and closed on the 26^th^ June 2020. Analysis was carried out in November 2020 following data cleaning and SAIL linkage. This involved comparison of means to demonstrate any differences between time points. Presentation of the outcomes give the confidence interval of the difference between groups.

## Results

The ‘HAPPEN At Home’ survey had 1333 responses, from 161 primary schools across Wales. Following the exclusion process presented in Figure 1 (no consent for linkage, missing FSM data), the final linked data (‘HAPPEN At Home’ responses and FSM status) for subsequent analysis included 574 participants.

**Figure 1.**
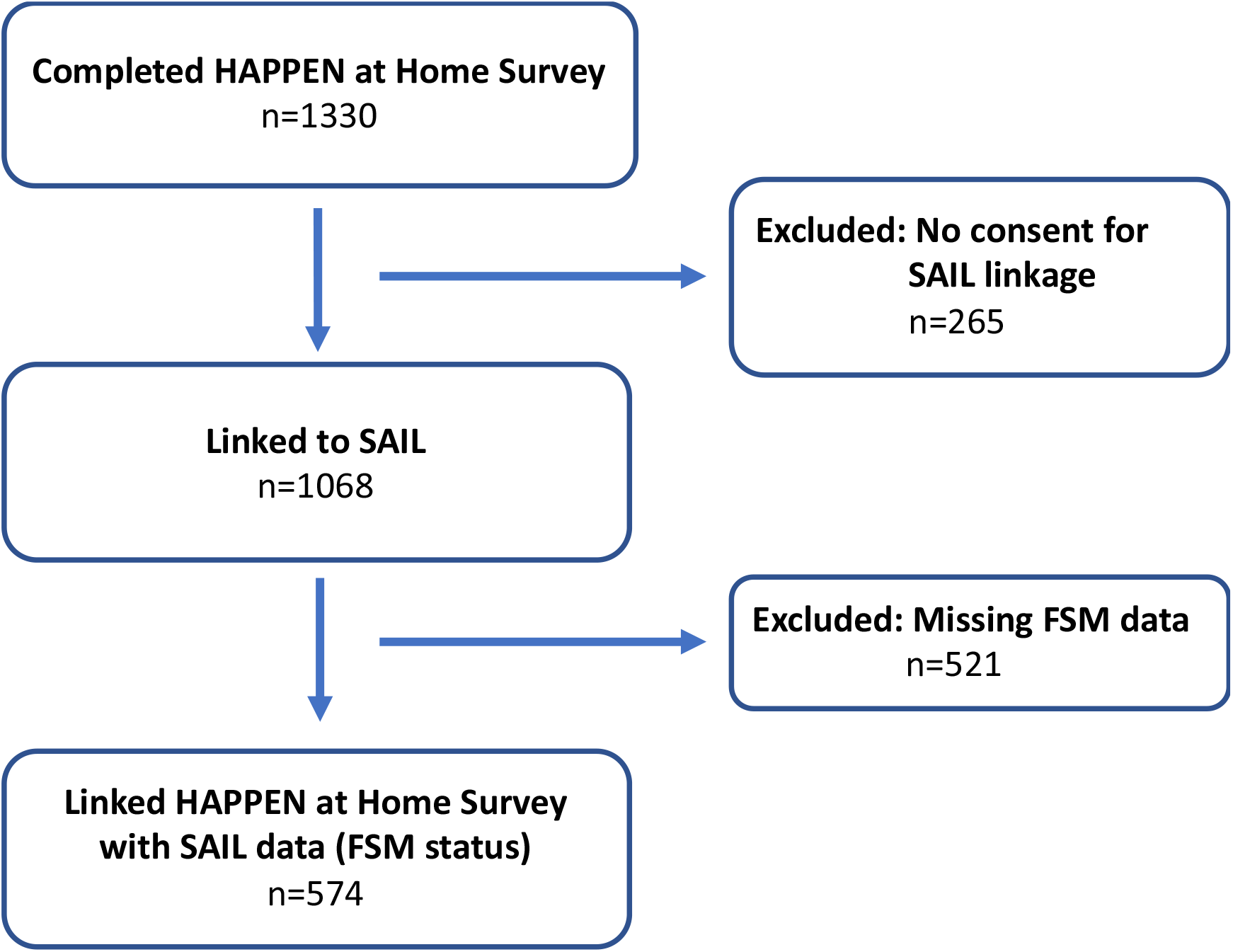
Study Flow Diagram

Data were stratified by FSM status and compared with 2019 from the same time period (March to June 2019). A breakdown of demographics by FSM status and time period is presented in Table 1.

**Table 1.** Demographics

| Demographics |  | March to June 2018 (n=475) | March to June 2019 (n=1150) | School closures 2020 (n=1068) | Difference (2019 – 2020) |
| --- | --- | --- | --- | --- | --- |
| Gender | Boy | 233 (49.19%) | 594 (51.65%) | 535 (50.09%) | -1.56% (-.26 to 5.71) |
|  | Girl | 241 (50.65%) | 548 (47.65%) | 528 (49.44%) | 1.79% (-2.37 to 5.94) |
|  | Prefer Not To Say | 1 (0.16%) | 8 (0.70%) | 5 (0.47%) | -.23% (-.04 to .09) |
| Age | Mean | 10.30 | 10.27 | 9.99 | -.28 (-.36 to -.19) |
| Year Group | 3 | NA | NA | 92 (8.61%) | NA |
|  | 4 | 69 (14.54%) | 303 (26.35%) | 373 (34.93%) | 8.58% (.47 to 12.39) |
|  | 5 | 233 (49.12%) | 403 (35.04%) | 283 (26.50%) | -8.54% (.47 to 12.34) |
|  | 6 | 173 (36.35%) | 444 (38.61%) | 320 (29.96%) | -8.65% (.47 to 12.55) |

### Differences in health outcomes before (2018-2019) and during school closures (2020)

During school closures there was a significant improvement in physical activity (see Table 2) (4.5% increase in number achieving 60 minutes of physical activity a day (95%CI: 0.95% to 8.14%) and in sleep (10.39% more having the recommended 9 hours sleep). Children also report feeling less tired. There were no significant differences in these variables between 2018 and 2019 suggesting that these findings are associated with lockdown restrictions and school closures as opposed to time trends. Perceptions of general competency and feeling safe in your area (S1) also increased during school closures.

**Table 2.**
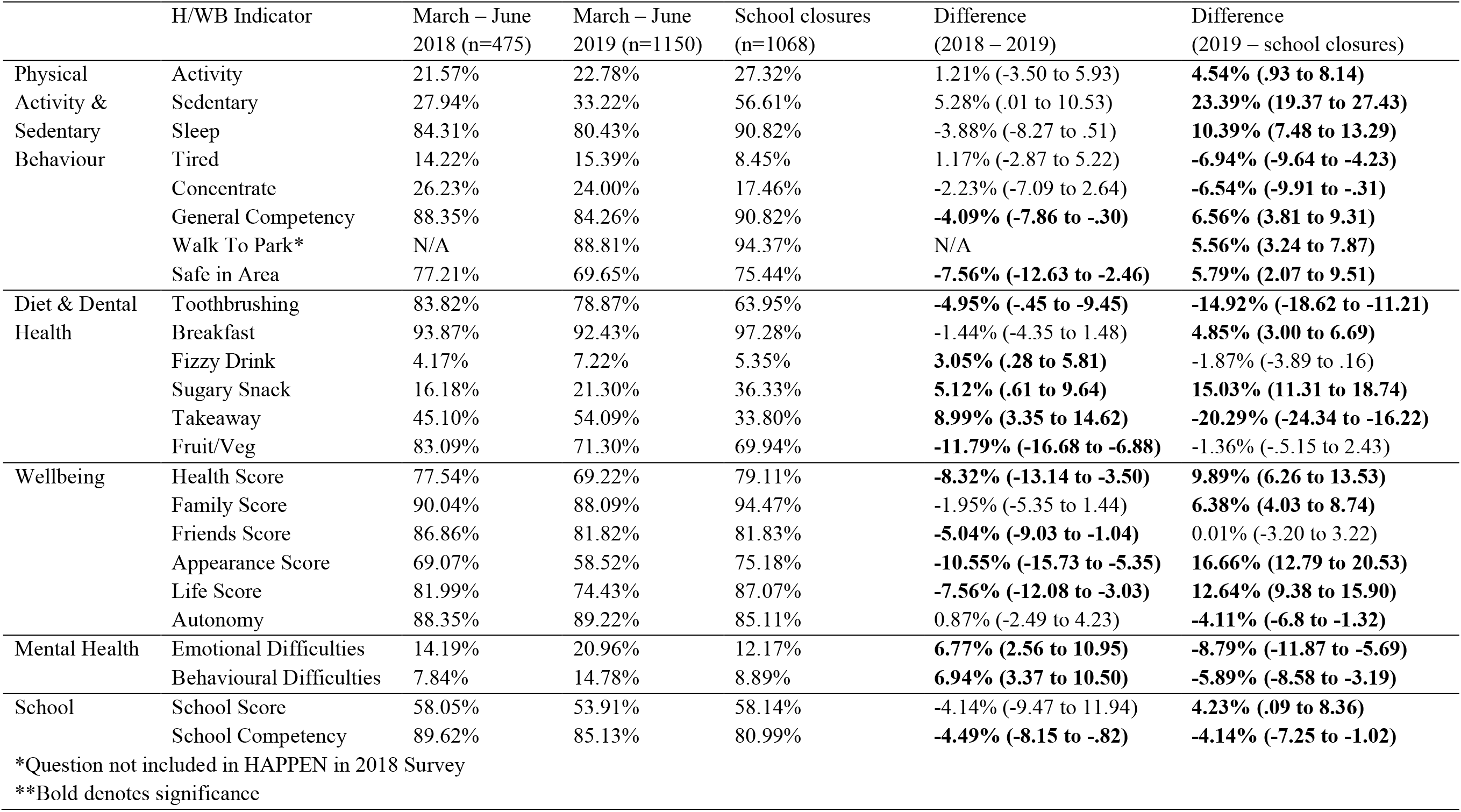
Differences between those who took part in the HAH survey and those who have taken part in HAPPEN previously (group comparison between 2018, 2019 and 2020)

Regarding dietary and dental health behaviours, the amount of daily teeth brushing decreases annually (Table 2) but this is more significant between 2019 and 2020 (−14.92%, CI: -18.62 to -11.21). Interestingly the number of takeaways consumed per week has significantly decreased during 2020 (−20.29%, CI: -24.34 to -16.33) while sugary snack consumption has increased (15.03%, CI: 11.31 to 18.74). However, there appears to be an annual trend in sugary snack consumption when compared to 2019 and 2018 data. A higher proportion of children report eating breakfast during school closures compared to previous years.

Between 2018 and 2019, wellbeing shows significant decreases in a number of areas including perceptions of health, friends, appearance and life. However, during school closures this trend reversed (table 2). Most notably children reported being happier with their health (9.89%, CI: 6.26 to 13.53), appearance (16.66%, CI: 12.79 to 20.53) and life (12.64%, CI: 9.38 to 15.90). A similar trend is evident in terms of mental health (fewer emotional and behavioural difficulties) (table 2).

Despite being away from the school environment, children report feeling happier with school. Yet their self-reported school competency was reduced during school closures. However, there is an annual decrease since 2018 suggesting a temporal trend in pupils’ perception of school ability.

### Differences in health outcomes before (2018-2019) and during school closures (2020) stratified by deprivation (FSM eligibility)

Compared to non-FSM children (Table 3), those eligible for FSM walked to the park less, their takeaway consumption showed less decline but their fruit and vegetable consumption significantly declined (−21.28%, CI: -37.08 to -5.67). This decline was not seen in non-FSM children. The decline in perceptions of school competency from 2019 to 2020 was three times higher within the FSM group.

**Table 3.** Exploring any differences between those who took part in the HAH survey and those who have taken part in 2019 stratified by FSM

|  |  | March – June 2019 |  | School closures 2020 |  |  |  |
| --- | --- | --- | --- | --- | --- | --- | --- |
| H/WB Indicator |  | No FSM<br>(n=520) | FSM<br>(n=120) | No FSM<br>(n=384) | FSM (n=53) | No FSM Difference | FSM Difference |
| Physical<br>Activity &<br>Sedentary<br>Behaviour | Activity | 25.00% | 23.33% | 28.12% | 15.09% | 3.12% (-2.69 to 8.94) | -8.24% (-21.47 to 4.99) |
|  | Sedentary | 29.61% | 42.50% | 60.67% | 58.49% | **31.06% (24.84 to 37.28)** | 15.99% (-0.18 to 32.16) |
|  | Sleep | 81.34% | 75.83% | 89.84% | 83.01% | **8.50% (3.80 to 13.19)** | 7.18% (-6.32 to 20.69) |
|  | Tired | 14.03% | 25.83% | 9.11% | 20.75% | **-4.92% (-9.19 to -0.64)** | -5.08% (-19.09 to 8.94) |
|  | Concentrate | 26.15% | 16.66% | 16.66% | 18.86% | **-9.49% (-14.93 to -4.03)** | 2.20% (-10.19 to 14.59) |
|  | General Competency | 85.38% | 78.33% | 92.18% | 90.56% | **6.80% (2.57 to 11.03)** | 12.23% (-0.18 to 24.65) |
|  | Walk To Park | 87.23% | 94.06% | 94.01% | 92.45% | **6.78% (2.85 to 10.70)** | -1.61% (-9.66 to 6.43) |
|  | Safe in Area | 72.11% | 54.16% | 76.82% | 62.26% | 4.71% (-10.74 to 10.48) | 8.10% (-8.08 to 24.27) |
| Diet & Dental<br>Health | Toothbrushing | 83.46% | 64.16% | 63.80% | 47.16% | **-19.65% (-25.22 to -14.08)** | **-17.00% (-32.89 to -1.09)** |
|  | Breakfast | 94.03% | 86.66% | 97.65% | 92.45% | **3.62% (0.90 to 6.32)** | 5.79% (-4.64 to 16.22) |
|  | Fizzy Drink | 5.76% | 13.33% | 4.68% | 7.54% | -1.08% (-4.04 to 1.88) | -5.79% (-16.22 to 4.64) |
|  | Sugary Snack | 20.38% | 18.33% | 36.97% | 32.07% | **16.59% (10.79 to 22.39)** | **13.74% (0.21 to 27.26)** |
|  | Takeaway | 54.23% | 57.50% | 34.89% | 50.94% | **-19.34% (-25.80 to -12.86)** | -6.56% (-22.80 to 9.68) |
|  | Fruit/Veg | 74.23% | 66.66% | 68.75% | 45.28% | -5.48% (-11.41 to 0.45) | **-21.38% (-37.08 to -5.67)** |
| Wellbeing | Health Score | 72.30% | 60.83% | 78.64% | 66.03% | **6.34% (0.62 to 12.04)** | 5.20% (-10.63 to 21.04) |
|  | Family Score | 89.03% | 81.66% | 94.01% | 92.45% | **4.98% (1.23 to 8.71)** | **10.79% (0.80 to 22.37)** |
|  | Friends Score | 83.07% | 72.50% | 83.33% | 71.69% | 0.26% (-4.68 to 5.20) | -0.81% (-15.46 to 13.85) |
|  | Appearance Score | 60.38% | 45.83% | 75.78% | 66.03% | **15.40% (9.25 to 21.53)** | **20.20% (4.13 to 36.27)** |
|  | Life Score | 74.80% | 63.33% | 89.32% | 79.24% | **14.52% (9.41 to 19.61)** | **15.91% (0.85 to 30.97)** |
|  | Autonomy | 88.65% | 88.33% | 85.41% | 84.90% | **-3.24% (-7.63 to 11.62)** | -3.43% (-14.32 to 7.46) |
| Mental Health | Emotional Difficulties | 18.65% | 26.66% | 10.93% | 20.75% | **-7.72% (-12.45 to -2.97)** | -5.91% (-20.03 to 8.21) |
|  | Behavioural Difficulties | 14.42% | 27.50% | 6.77% | 20.75% | **-7.65% (-11.78 to -3.51)** | -6.75% (-20.96 to 7.47) |
| School | School Score | 66.34% | 61.66% | 74.47% | 56.60% | **8.13% (2.08 to 14.17)** | -5.06% (-21.07 to 10.95) |
|  | School Competency | 86.34% | 80.00% | 79.42% | 58.49% | **-6.92% (-11.81 to -2.02)** | **-21.51% (-35.60 to -7.41)** |

\*Bold denotes significance

During school closures, there was a significant difference of reported daily physical activity between those on FSM and those not on FSM (13.03% difference, 95% CI: 3.3% to 21.66%). Compared to non-FSM children, a lower proportion of FSM eligible children reported to engage in at least 60 minutes of daily physical activity during school closures (non-FSM: 28.12%; FSM: 15.09%). Children not on FSM showed a significant increase in sedentary time and reported a lower ability to concentrate. This was not a significant trend notices for those on FSM. However, there was an increase from 2019 to 2020 in family wellbeing scores for all children and especially among those eligible for FSM.

## Discussion

Improvements during school closures for children included physical activity, sleep, wellbeing (family, health, life) and emotional and behavioural difficulties when considering the group as a whole. Primary school children also report higher wellbeing especially family score, during lockdown. However, aspects which were detrimental during school closures included less tooth brushing for all children. FSM children reported a reduction in the time spent engaged in physical activity, significantly less fruit and vegetable consumption and lower self-assessed school competence than before school closures.

### Physical activity and sedentary behaviour

Overall, small improvements to time spent being physically active were seen during school closures. However, this increase is likely to be amongst non-FSM pupils. For those on FSM activity decreased and may be due to less access to safe areas to play compared to those not on FSM.

Recent research around school staff perceptions of the return to school echo this finding. Teachers perceived that their pupils had been less active during lockdown restrictions and observed upon the phased return to school that some children had gained weight^21^. Findings from the current study suggest this may be more pronounced for more deprived pupils. Those eligible for FSM felt less safe in their areas which may be why they were less active.

Non-FSM children were more active. However, non-FSM children’s sedentary time was significantly higher during school closures. Their reported daily screen time (>2 hours) doubled compared to the previous year. The delivery of education during school closures was achieved primarily online through home learning and thus, children will have utilized screens (e.g., laptops and tablets) to aid learning. Less deprived families may have better access to these resources and therefore, screen time may be higher in this group. This is supported by research from the Institute for Fiscal Studies^22^ where children from less deprived families were spending 30% more time engaging in home learning activities than those more deprived. This may also reflect why perceptions of school competency remains much higher in the less deprived group. This suggests that non-FSM children were more engaged with learning tasks and therefore had perceived higher competence and confidence with learning and development. This may contribute towards the estimated 46% increase in learning gap between disadvantaged children and their peers reported by teachers^23^. With the relationship between education and health well documented, this has implications for children’s future health and wellbeing outcomes^24^. Further evidence of this is seen in feeling part of your school community which again is much higher in those not on FSM.

For those eligible for FSM, the amount of sedentary time may appear positive in comparison to non-FSM but could also highlight inequalities relating to digital poverty and contribute to gaps in learning progression. Previous HAPPEN research^21^ has highlighted the lack of access to digital equipment, sharing devices and a lack of digital competency in accessing home learning. This is worth noting as while less screen time could be perceived as a benefit to physical health in FSM children, during school closures it could also mean that learning gaps are being widened.

Children not on FSM report to not being able to concentrate as much compared to the previous year. The increased sedentary time may be due to increased screen time/online working for non-FSM pupils during school closures may have been detrimental to concentration. More research is needed into how screen time was consumed during school closures and the impact this has on health is required.

### Diet and dental health

Toothbrushing was significantly lower in children compared to 2019 regardless of FSM status. This meant many children were brushing their teeth less than the recommended guidelines of twice per day. Research shows that lack of routine and structure puts children at risk of poorer dental hygiene^25^ which can have long-term impacts. It is possible that school closures disrupted bedtime and wake time routines in which teeth brushing would usually take place, and therefore may account for the lower frequency of teeth brushing. In addition, the lack of access to school-based dental hygiene programmes such as ‘Designed to Smile’^26^ may have a significant impact on teeth brushing behaviour. This coupled with observed increases in sugary snack consumption through school closures may have a detrimental impact on dental hygiene.

Those on FSM saw the biggest impact on dietary behaviours during lockdown restrictions. Not only was takeaway consumption higher in this group, but FSM children also consumed fewer fruit and vegetables during this time. FSM are a key public health policy to aid in reducing food insecurity and associated negative health and educational inequalities in the UK. It appears that those utilising FSM have been significantly impacted by school closures in not being able to access regular meal provision in a school setting in Wales. Research shows that almost half of all children on FSM were unable to access them during school closures^27^.

Providing children with nutritional meals in school helps to narrow health inequalities and the educational attainment gap between the most and least deprived children^29,30^. Findings from this study add further evidence to disparities amongst groups of children from different backgrounds. While the initial lockdown in March 2020 was temporary, the findings of the current study support the mounting evidence that prolonged lockdown periods will affect children’s physical health^27^.

### Wellbeing and mental health

Within this current study, improvements in family wellbeing was observed during school closures for both groups of children. This is likely due to an increased number of parents working from home or being furloughed, enabling some children to spend extra time that they otherwise would not have had with caregivers. School staff acknowledge this, they reported children having more opportunities for walking, exploring and spending time outside, with this contributing to strengthened family relationships^21^.

Happiness with life was also significantly higher generally and increased equally in both groups from 2019 data. It is important to note that deprived children still report feeling less happy in general compared to non-FSM children. The findings regarding physical activity may underpin this, with increased opportunities to play and be outdoors, for example having more time during lockdown and feeling safer in their areas. Moreover, behavioural and emotional difficulties reported during school closures was significantly lower. In less deprived children, this number was almost half suggesting a more positive impact in those not on FSM. Interestingly, previous research has found the opposite, with parents and teachers reporting increases in emotional and behavioural difficulties as well as low mood, anxiety and social disconnection^21,31^. It is possible these conflicting findings highlight the difference between child reported and adult external observations.

## Limitations

Although the ‘HAPPEN At Home’ survey was made available to all children aged 8-11 across Wales, the findings of this paper only present those who participated in the survey and a subsample who consented to data linkage. As the survey took place at home due to school closures, those who participated will be from families who have internet access. The difference in inequalities are likely to be much higher among those who could not participate due to lack of access to the internet. While the sampling strategy was the same for 2020 and 2019, 2018 data was sampled more purposefully from South Wales which may have an influence on findings from this year.

There is evidence that FSM status is not a perfect measure of socio-economic deprivation ^32^ and there are also a number of other factors that contribute to the deprivation levels of a child. However, FSM status does come very close to identifying a group of children who may be at disadvantage due to their socio-economic position^32^.

## Conclusion

Overall, findings from this study show that, as a group, many things improved during the period of school closures for children including physical activity, sleep and general wellbeing. However, there are significant differences and inequalities when stratified by FSM. Improvements were mostly observed in non-FSM children. For children eligible for FSM, diet (e.g. lower fruit and vegetable intake), physical activity and dental health was significantly impacted. These findings are concerning as they illustrate the importance of the entire school day, including free school meal provision, in attenuating physical health inequalities in children.

This paper shows the short-term effect of school closures on children’s health and wellbeing. Furthermore, this research highlights a number of concerns regarding wider physical health inequalities such as obesity. When schools reopen this research suggests there will be a need to address wider physical health inequalities such as obesity, poor dental health, lack of vitamins and minerals and lower fitness in those from deprived backgrounds.

## Data Availability

Data collected for this study, including individual participant data and a data dictionary will not be made available.

## Author contributions

MJ wrote the first draft of the paper and all authors provided critical input and revisions for all further drafts. MJ, EM and SB designed data collection and MJ and SB undertook data analysis. MJ, EM, SB, MD and JW aided in interpretation of findings and supervision of study quality. The corresponding author attests that all listed authors meet authorship criteria and that no others meeting the criteria have been omitted.

