## Supplementary Information for "Impact of school closures on the health and well-being of primary school children in Wales UK; a routine data linkage study using the HAPPEN survey (2018-2020)"

**S1**
**Items included in the analyses**

| Health and wellbeing topic | Item within HAPPEN at Home Survey |
|---|---|
| Physical activity and sedentary behaviour | *"In the last 7 days, how many days did you do sports or exercise for at least 1 hour in total. This includes doing any activities (including online activities like Joe Wicks) or playing sports where your heart beat faster, you breathed faster and you felt warmer?" (e.g. 5-6 days)*<br><br>*"In the last 7 days, how many days did you watch TV/play online games/use the internet etc. for 2 or more hours a day (in total)?"*<br><br>*"What time did you wake up TODAY (to the nearest half hour)?"*<br><br>*"On a scale of 0 to 10 (0 being not very safe and 10 being very safe), how safe do you feel playing in your area?"* |
| Diet and dental health | *"How many times did you brush your teeth YESTERDAY?"*<br><br>*"In the last 7 days, how many days did you drink at least one fizzy drink (e.g. coke, fanta, sprite)?"*<br><br>*"Did you eat any fruit and vegetables YESTERDAY?"* |
| Wellbeing | *"On a scale of 0 to 10 (0 being very unhappy and 10 being very happy), how do you feel about:*<br><br>*Your Health?*<br>*Your Family?*<br>*Your Friends?*<br>*Your Appearance?*<br>*Your Life?"*<br><br>**From the Good Childhood Index (2010) developed by the Children's Society* |
| Mental health | *"Remember, there are no right or wrong answers, just pick which is right for you.*<br><br>*I feel lonely.*<br>*I cry a lot.*<br>*I am unhappy"*<br><br>**From the Me and My Feelings Questionnaire* |
| School | *"On a scale of 0 to 10 (0 being very unhappy and 10 being very happy), how do you feel about:*<br><br>*Your School?"*<br><br>**From the Good Childhood Index (2010) developed by the Children's Society* |

| | |
|---|---|
| | *"Tell us if you agree or disagree with the following:*<br><br>*I am doing well with my school work" (e.g. Strongly agree, agree, don't agree or disagree, disagree, strongly disagree)* |

### THE HAPPEN SURVEY [At Home]

* Required

#### Consent Form

**Before you start please click this link to read the information sheet...**
https://happen-wales.co.uk/childrens-information-sheet/

1. I have read the child information sheet and understand that if I take part I can change my mind at any time, and this will not be a problem at all. *

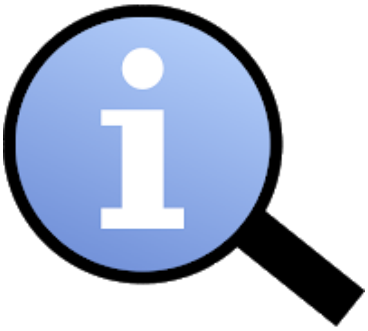

*Mark only one oval.*

- ◯ Yes
- ◯ No

2. I am happy for you to use my questionnaire for research. Only the researchers in the team will know my name and will not tell anyone else my answers *

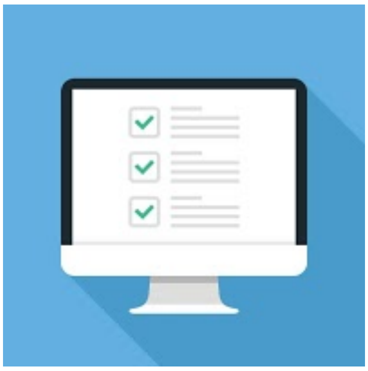

*Mark only one oval.*

- ⬭ Yes
- ⬭ No do not use my questionnaire

3. I am happy for you to look at my school and health records to see how my school is doing (as a group). This is anonymous which means I cannot be identified *

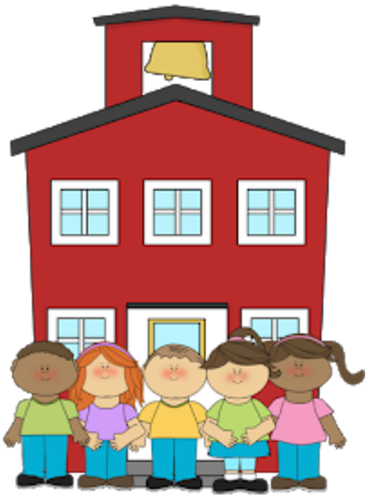

*Mark only one oval.*

- ⬭ Yes
- ⬭ No

If you do not wish to take part in the questionnaire please do not continue.

Please click next to start the questionnaire!

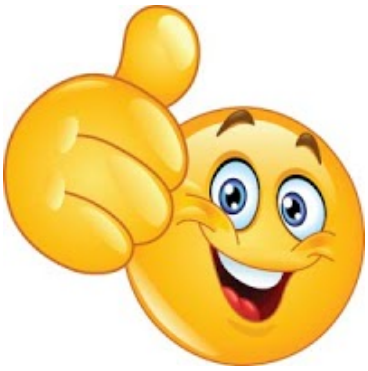

About You

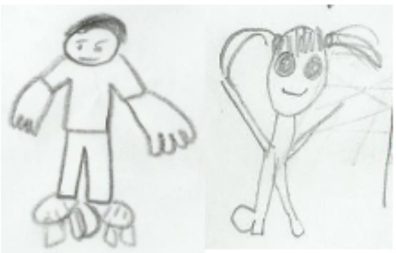

4. First Name *

5. Last Name *

6. Home Post Code *

7. What school do you go to? *

8. Are you still going to school? *

*Mark only one oval.*

- No, I am at home
- Yes, most days of the week
- Yes, sometimes
- I am in a different school from my own school

9. Do you have any other children living in your house with you? *

*Mark only one oval.*

- Yes
- No

10. How many people live in your home with you (including adults)? *

*Mark only one oval.*

- 1
- 2
- 3
- 4
- 5
- 6+

11. What year are you in now? *

*Mark only one oval.*

- Year 4
- Year 5
- Year 6

12. Gender *

*Mark only one oval.*

- Boy
- Girl
- Prefer not to say

#### Date of Birth

13. Year *

*Mark only one oval.*

- 2007
- 2008
- 2009
- 2010
- 2011
- 2012

14. Month *

*Mark only one oval.*

- January
- February
- March
- April
- May
- June
- July
- August
- September
- October
- November
- December

15. Day *

*Mark only one oval.*

- 1
- 2
- 3
- 4
- 5
- 6
- 7
- 8
- 9
- 10
- 11
- 12
- 13
- 14
- 15
- 16
- 17
- 18
- 19
- 20
- 21
- 22
- 23
- 24
- 25
- 26
- 27
- 28
- 29
- 30
- 31

YESTERDAY

### CHAT

Child Health and Activity Tool

Firstly, think carefully about what you did <u>YESTERDAY</u> and then answer the following questions....

16. 1. What did you eat for breakfast YESTERDAY? *

*Check all that apply.*

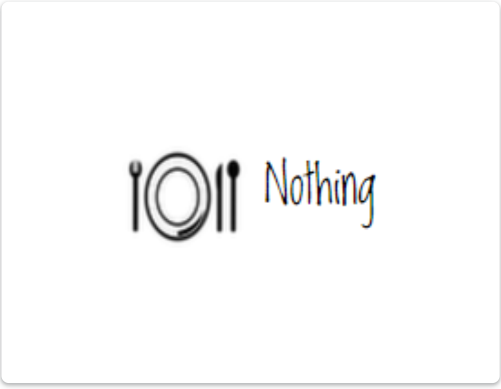


- [ ] Nothing

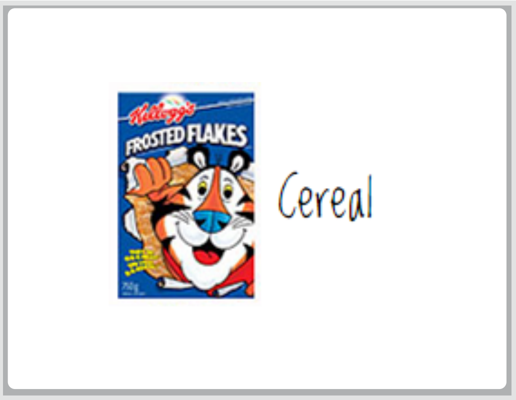


- [ ] Sugary cereal e.g. cocopops, frosties, sugar puffs, chocolate cereals

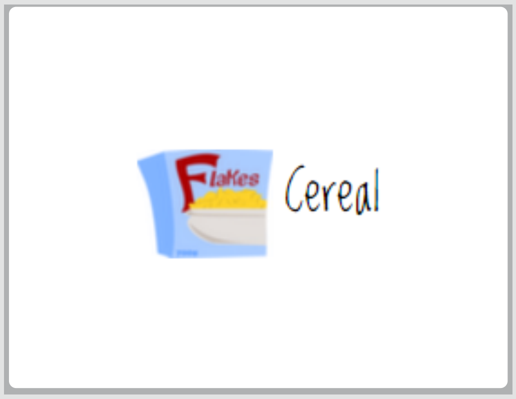


- [ ] Healthy cereal e.g. porridge, weetabix, readybrek, muesli, branflakes, cornflakes

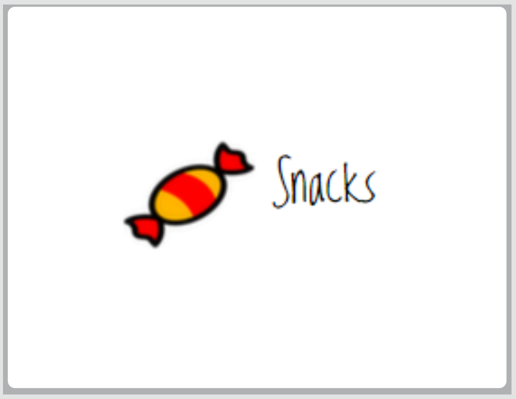


- [ ] Snacks

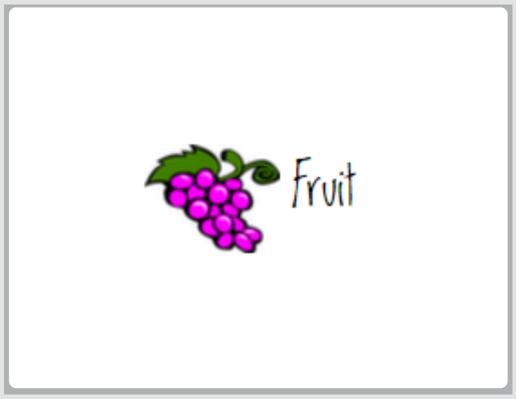


- [ ] Fruit

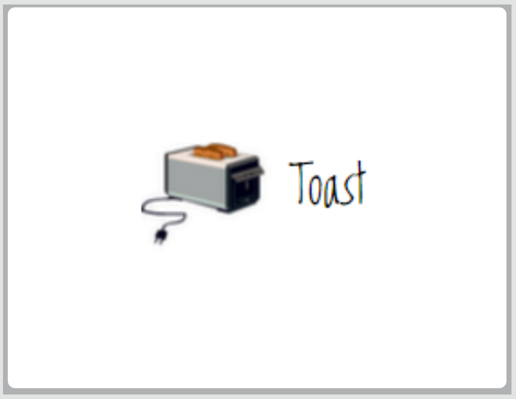


- [ ] Toast

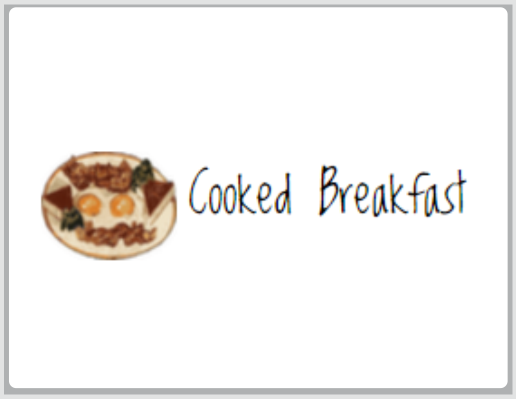


- [ ] Cooked breakfast

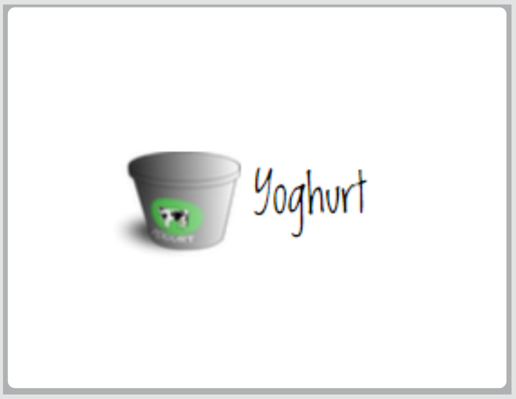


- [ ] Yoghurt

Other: ☐ ______________

17. 2. Did you eat any fruit and vegetables YESTERDAY? *

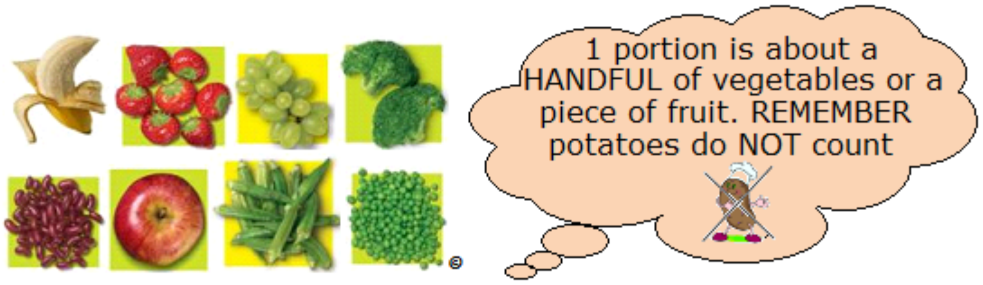


*Mark only one oval.*

- ⬭ No
- ⬭ 1 Piece
- ⬭ 2 Or More Fruit and Veg

18. 3. How many times did you brush your teeth YESTERDAY? *

*Mark only one oval.*

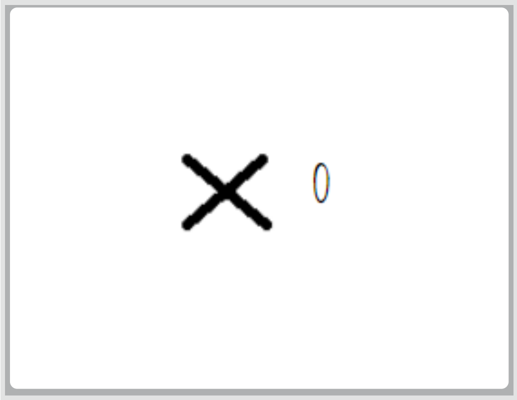


⬭ 0

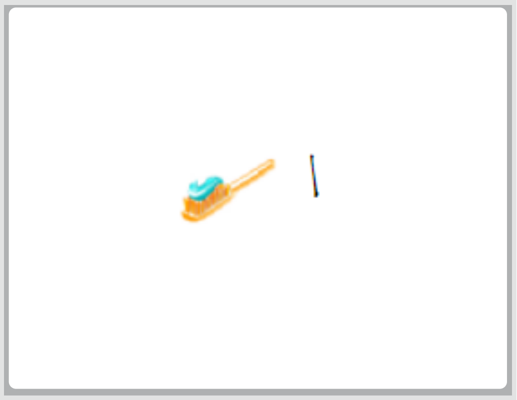


⬭ 1

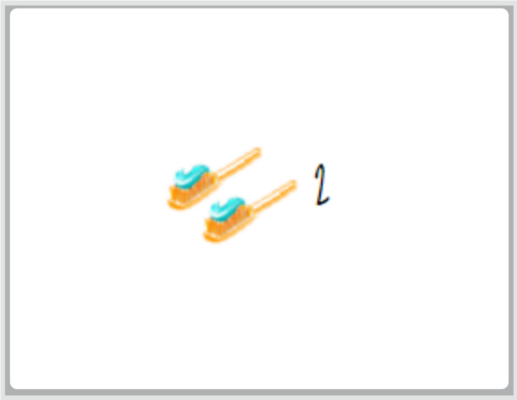


⬭ 2

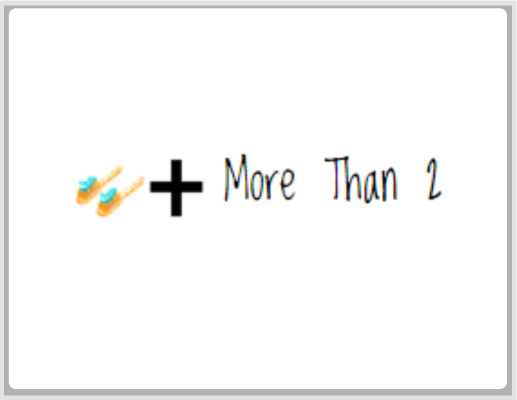


⬭ 3

19. 4. What time did you fall asleep YESTERDAY (to the nearest half hour)? *

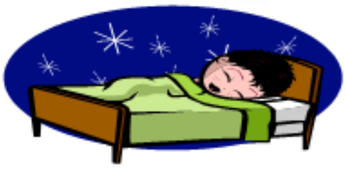

*Mark only one oval.*

- 6:00pm
- 6:30pm
- 7:00pm
- 7:30pm
- 8:00pm
- 8:30pm
- 9:00pm
- 9:30pm
- 10:00pm
- 10:30pm
- 11:00pm
- 11:30pm
- 12:00am
- 12:30am
- 1:00am
- 1:30am
- 2:00am
- 3:00am
- 3:30am
- 4:00am

20. 5. What time did you wake up TODAY (to the nearest half hour)? *

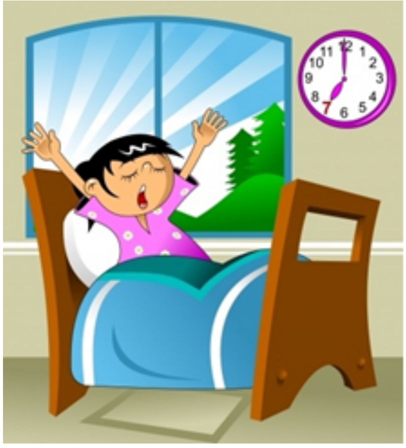

*Mark only one oval.*

- ◯ 5:00am
- ◯ 5:30am
- ◯ 6:00am
- ◯ 6:30am
- ◯ 7:00am
- ◯ 7:30am
- ◯ 8:00am
- ◯ 8:30am
- ◯ 9:00am
- ◯ 9:30am
- ◯ 10:00am
- ◯ 10:30am
- ◯ 11:00am
- ◯ 11:30am

THE LAST WEEK

NOW think about what you did in the last 7 days...

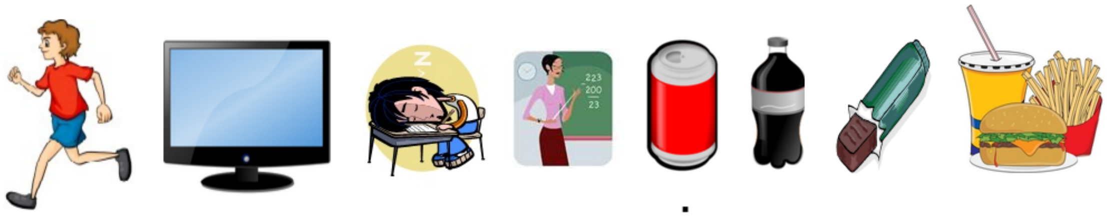

21. 6. In the last 7 days, how many days did you do sports or exercise for at least 1 hour in total. This includes doing any activities (including online activities like Joe Wicks) or playing sports where your heart beat faster, you breathed faster and you felt warmer? *

*Mark only one oval.*

- 0 days
- 1-2 days
- 3-4 days
- 5-6 days
- 7 days

22. 7. In the last 7 days, how many days did you watch TV/play online games/use the internet etc. for 2 or more hours a day (in total)? *

*Mark only one oval.*

- 0 days
- 1-2 days
- 3-4 days
- 5-6 days
- 7 days

23. 8. In the last 7 days, how many days did you feel tired? *

*Mark only one oval.*

- 0 days
- 1-2 days
- 3-4 days
- 5-6 days
- 7 days

24. 9. In the last 7 days, how many days did you feel like you could concentrate/pay attention well on your school work? *

*Mark only one oval.*

- 0 days
- 1-2 days
- 3-4 days
- 5-6 days
- 7 days

25. 10. In the last 7 days, how many days did you drink at least one fizzy drink (e.g. coke, fanta, sprite)? *

*Mark only one oval.*

- 0 days
- 1-2 days
- 3-4 days
- 5-6 days
- 7 days

26. 11. In the last 7 days, how many days did you eat at least one sugary snack (e.g. chocolate bar, sweets)? *

*Mark only one oval.*

- 0 days
- 1-2 days
- 3-4 days
- 5-6 days
- 7 days

27. 12. In the last 7 days, how many days did you eat take away foods (e.g. Chinese takeaway)? *

*Mark only one oval.*

- ⬭ 0 days
- ⬭ 1-2 days
- ⬭ 3-4 days
- ⬭ 5-6 days
- ⬭ 7 days

Activity and Your Area

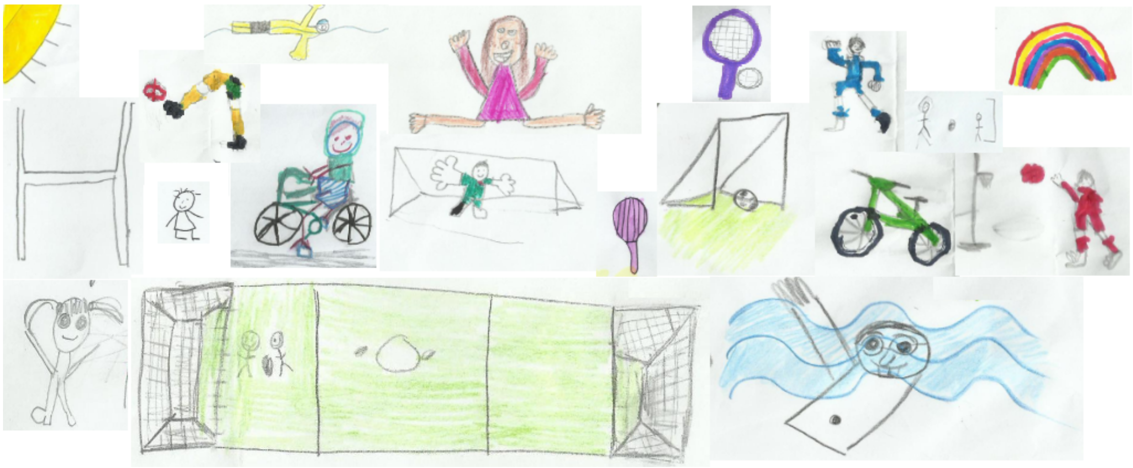

28. 13. On a scale of 0 to 10 (0 being not very safe and 10 being very safe), how safe do you feel playing in your area?

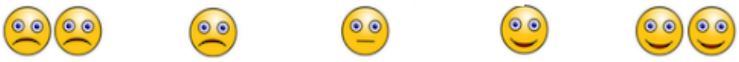

*Mark only one oval.*

| | 0 | 1 | 2 | 3 | 4 | 5 | 6 | 7 | 8 | 9 | 10 | |
|---|---|---|---|---|---|---|---|---|---|---|---|---|
| Not very safe | ⬭ | ⬭ | ⬭ | ⬭ | ⬭ | ⬭ | ⬭ | ⬭ | ⬭ | ⬭ | ⬭ | Very safe |

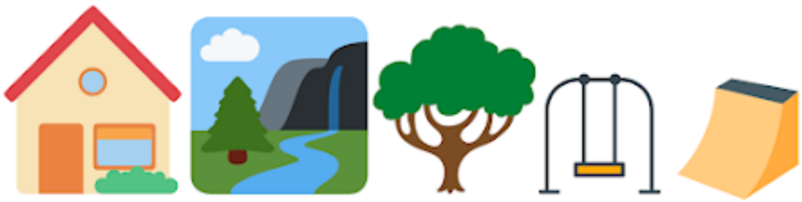

29. 14. From your house, can you easily walk to a park (for example a field or grassy area)? *

*Mark only one oval.*

- Yes
- No

30. 15. From your house, can you easily walk to somewhere you can play? *

*Mark only one oval.*

- Yes
- No

31. 16. Do you have a garden? *

*Mark only one oval.*

- Yes
- No

32. 17. How often do you go out to play outside? *

*Mark only one oval.*

- Most days
- A few days each week
- Hardly ever
- I don't play

33. 18. Do you have enough time for play? *

*Mark only one oval.*

- ( ) Yes, I have loads
- ( ) Yes, it's just about enough
- ( ) No, I would like to have a bit more
- ( ) No, I need a lot more

34. 19. What type of places do you play in? *

*Check all that apply.*

- [ ] In my house
- [ ] In my garden
- [ ] In the street
- [ ] On a local grassy area
- [ ] In a place with bushes, trees and flowers
- [ ] In the woods near my house
- [ ] On a football field near my house
- [ ] In my school playground
- [ ] Somewhere with water or sand in it
- [ ] On the bike or skate park

Other: [ ] ______________________

35. 20. Can you play in all the places you would like to? *

*Mark only one oval.*

- ( ) I can play in all the places I would like to
- ( ) I can play in some of the places I would like to
- ( ) I can only play in a few places I would like to
- ( ) I can hardly play in any of the places I would like to

36. 21. Do you have somewhere at home where you have space to relax? *

*Mark only one oval.*

- ( ) Yes
- ( ) Sometimes but not all the time
- ( ) No

You and Your Feelings

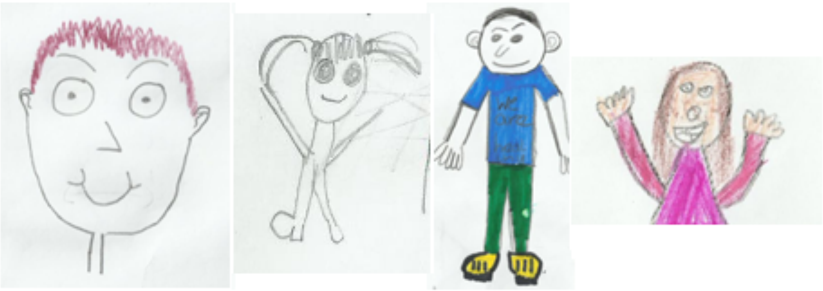

This part of the survey is going to ask you how you feel. There are no right or wrong answers. You should just pick the answer which is best for you.

37. 22. Tell us if you agree or disagree with the following: *

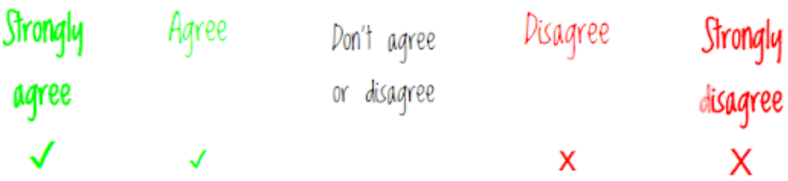


*Mark only one oval per row.*

| | Strongly agree | Agree | Don't agree or disagree | Disagree | Strongly disagree |
|---|---|---|---|---|---|
| I am doing well with my school work | | | | | |
| I feel part of my school community | | | | | |
| I have lots of choice over things that are important to me | | | | | |
| There are lots of things I'm good at | | | | | |

23. On a scale of 0 to 10 (0 being very unhappy and 10 being very happy), how do you feel about:

38. Your Health *

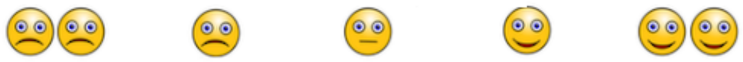

*Mark only one oval.*

| | 0 | 1 | 2 | 3 | 4 | 5 | 6 | 7 | 8 | 9 | 10 | |
|---|---|---|---|---|---|---|---|---|---|---|---|---|
| Very unhappy | ◯ | ◯ | ◯ | ◯ | ◯ | ◯ | ◯ | ◯ | ◯ | ◯ | ◯ | Very happy |

39. Your School (Not Home School) *

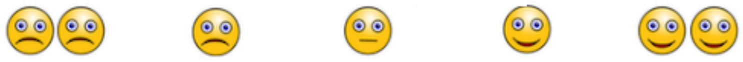

*Mark only one oval.*

| | 0 | 1 | 2 | 3 | 4 | 5 | 6 | 7 | 8 | 9 | 10 | |
|---|---|---|---|---|---|---|---|---|---|---|---|---|
| Very unhappy | ◯ | ◯ | ◯ | ◯ | ◯ | ◯ | ◯ | ◯ | ◯ | ◯ | ◯ | Very happy |

40. Your Family *

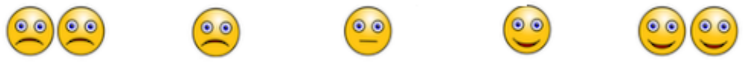

*Mark only one oval.*

| | 0 | 1 | 2 | 3 | 4 | 5 | 6 | 7 | 8 | 9 | 10 | |
|---|---|---|---|---|---|---|---|---|---|---|---|---|
| Very unhappy | ◯ | ◯ | ◯ | ◯ | ◯ | ◯ | ◯ | ◯ | ◯ | ◯ | ◯ | Very happy |

41. Your Friends *

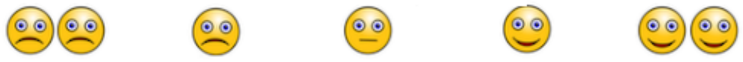

*Mark only one oval.*

| | 0 | 1 | 2 | 3 | 4 | 5 | 6 | 7 | 8 | 9 | 10 | |
|---|---|---|---|---|---|---|---|---|---|---|---|---|
| Very unhappy | ◯ | ◯ | ◯ | ◯ | ◯ | ◯ | ◯ | ◯ | ◯ | ◯ | ◯ | Very happy |

42. Your Appearance (how you look) *

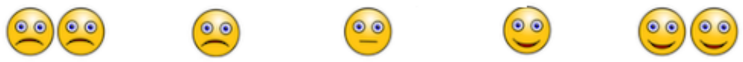

*Mark only one oval.*

| | 0 | 1 | 2 | 3 | 4 | 5 | 6 | 7 | 8 | 9 | 10 | |
|---|---|---|---|---|---|---|---|---|---|---|---|---|
| Very unhappy | ⬭ | ⬭ | ⬭ | ⬭ | ⬭ | ⬭ | ⬭ | ⬭ | ⬭ | ⬭ | ⬭ | Very happy |

43. Your Life *

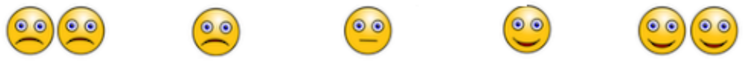

*Mark only one oval.*

| | 0 | 1 | 2 | 3 | 4 | 5 | 6 | 7 | 8 | 9 | 10 | |
|---|---|---|---|---|---|---|---|---|---|---|---|---|
| Very unhappy | ⬭ | ⬭ | ⬭ | ⬭ | ⬭ | ⬭ | ⬭ | ⬭ | ⬭ | ⬭ | ⬭ | Very happy |

#### You and Your Feelings

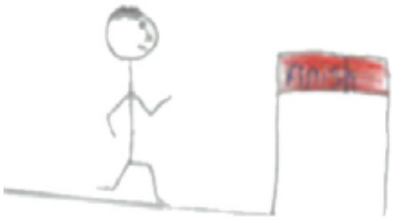

44. 24. Remember, there are no right or wrong answers, just pick which is right for you. *

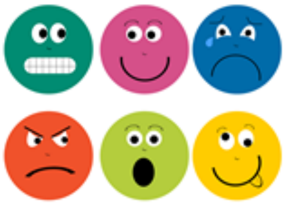

*Mark only one oval per row.*

| | Never | Sometimes | Always |
|---|---|---|---|
| I feel lonely | ⬭ | ⬭ | ⬭ |
| I cry a lot | ⬭ | ⬭ | ⬭ |
| I am unhappy | ⬭ | ⬭ | ⬭ |
| I feel nobody likes me | ⬭ | ⬭ | ⬭ |
| I worry a lot | ⬭ | ⬭ | ⬭ |
| I have problems sleeping | ⬭ | ⬭ | ⬭ |
| I wake up in the night | ⬭ | ⬭ | ⬭ |
| I am shy | ⬭ | ⬭ | ⬭ |
| I feel scared | ⬭ | ⬭ | ⬭ |
| I worry when I am at school | ⬭ | ⬭ | ⬭ |
| I get very angry | ⬭ | ⬭ | ⬭ |
| I lose my temper | ⬭ | ⬭ | ⬭ |
| I hit out when I am angry | ⬭ | ⬭ | ⬭ |
| I do things to hurt people | ⬭ | ⬭ | ⬭ |
| I am calm | ⬭ | ⬭ | ⬭ |
| I break things on purpose | ⬭ | ⬭ | ⬭ |

45. 25. Are you able to keep in touch with your family that you don't live with? *

*Mark only one oval.*

- ◯ Yes
- ◯ No

46. 20. Are you able to keep in touch with your friends? *

*Mark only one oval.*

- ◯ Yes
- ◯ No

47. 26. If yes, how are you keeping in touch (tick all you use)?

*Check all that apply.*

- ☐ I live near them so I can see them (at a social distance)
- ☐ By phone (texting, calling or video calling)
- ☐ On social media
- ☐ On games consoles

Thank you!

Well done, you've completed the questionnaire.

Thank you!

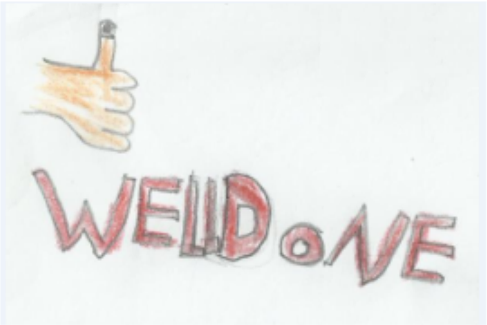

#### Don't forget to press submit below! Once you've pressed submit you are all done!

If you'd like some additional resources while you're at home during COVID-19, we've put together some here:

https://happen-wales.co.uk/some-resources-for-you/

### The HAPPEN Survey 2020/2021

* Required

#### Consent Form

Before you start please click this link to read the information sheet -> https://happen-wales.co.uk/childrens-information-sheet/ (https://happen-wales.co.uk/childrens-information-sheet/)

1

I have read the child information sheet -> https://happen-wales.co.uk/childrens-information-sheet/ (https://happen-wales.co.uk/childrens-information-sheet/) (click the link if you haven't read it) and understand that if I take part I can change my mind at any time, and this will not be a problem at all. *

◯ Yes

◯ No

2

I am happy for you to use my questionnaire for research. Only the researchers in the team will know my name and will not tell anyone else my answers. *

◯ Yes

◯ No do not use my questionnaire

3

I am happy for you to look at my school and health records to see how my school is doing (as a group). This is anonymous which means I cannot be identified. *

◯ Yes

◯ No

### About You

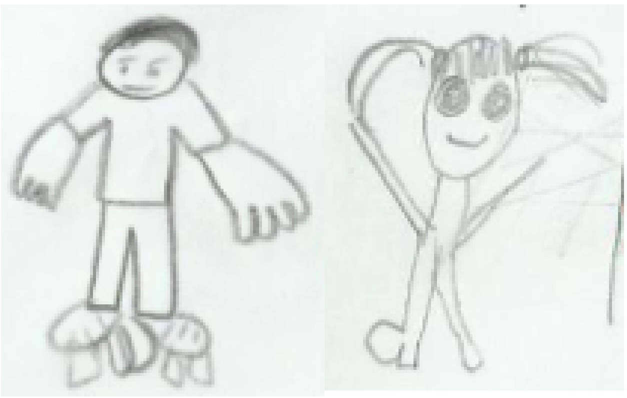

4

First Name

5

Last Name

6

Home Post Code

7

What school do you go to?

8

Do you have any other children living in your house with you?

9

What year are you in?

- ◯ Year 4
- ◯ Year 5
- ◯ Year 6

10

Do you have a garden?

- ◯ Yes
- ◯ No

11

Gender

- ◯ Boy
- ◯ Girl
- ◯ Prefer not to say

12

Are you...

- ◯ Asian
- ◯ Black
- ◯ White
- ◯ Mixed
- ◯ Prefer not to say

13

Date of Birth: Year

- ◯ 2007
- ◯ 2008
- ◯ 2009
- ◯ 2010
- ◯ 2011
- ◯ 2012

14

Date of Birth: Month

- ◯ January
- ◯ February
- ◯ March
- ◯ April
- ◯ May
- ◯ June
- ◯ July
- ◯ August
- ◯ September
- ◯ October
- ◯ November
- ◯ December

15

Date of Birth: Day

- ◯ 1
- ◯ 2
- ◯ 3
- ◯ 4
- ◯ 5
- ◯ 6
- ◯ 7
- ◯ 8
- ◯ 9
- ◯ 10
- ◯ 11
- ◯ 12
- ◯ 13
- ◯ 14
- ◯ 15
- ◯ 16
- ◯ 17
- ◯ 18
- ◯ 19
- ◯ 20
- ◯ 21
- ◯ 22
- ◯ 23

- ○ 25
- ○ 26
- ○ 27
- ○ 28
- ○ 29
- ○ 30
- ○ 31

Yesterday

Firstly, think carefully about what you did YESTERDAY and then answer the following questions....

16

How did you get to school YESTERDAY?

- ◯ On the bus
- ◯ On bike
- ◯ In the car/taxi
- ◯ Walked
- ◯ Ran/jogged
- ◯ Scooter
- ◯ Skateboarded/Rollerbladed

17

What did you have to eat for lunch YESTERDAY?

- ◯ School dinner
- ◯ Packed lunch from home
- ◯ Nothing

18

What did you do for MOST of your break-times YESTERDAY? (This includes lunchtime)

- ◯ Sat around inside or outside
- ◯ Ran around
- ◯ Stood around
- ◯ Walked around

19

How many friends did you play with?

- ◯ I like to play on my own
- ◯ 1-2
- ◯ 3-4
- ◯ 5 or more

20

Did you have an afternoon break at school?

- ◯ YES
- ◯ NO

21

How did you get home YESTERDAY?

- ◯ On the bus
- ◯ On bike
- ◯ In the car/taxi
- ◯ Walked
- ◯ Ran/jogged
- ◯ Scooter
- ◯ Skateboarded/Rollerbladed

After School

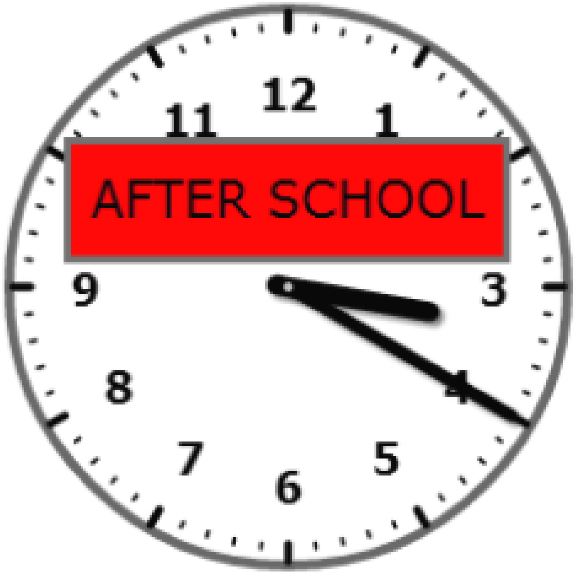


22

How many portions of fruit and vegetables did you eat YESTERDAY?

- ◯ 1
- ◯ 2
- ◯ 3
- ◯ 4
- ◯ 5
- ◯ 6
- ◯ 7
- ◯ 8

23

How many times did you brush your teeth YESTERDAY?

- ◯ 0
- ◯ 1
- ◯ 2
- ◯ 3

What time did you fall asleep YESTERDAY (to the nearest half hour)?

- ◯ 7:00pm
- ◯ 7:30pm
- ◯ 8:00pm
- ◯ 8:30pm
- ◯ 9:00pm
- ◯ 9:30pm
- ◯ 10:00pm
- ◯ 10:30pm
- ◯ 11:00pm
- ◯ 11:30pm
- ◯ 12:00am
- ◯ 12:30am
- ◯ 1:00am
- ◯ 1:30am
- ◯ 2:00am
- ◯ 2:30am
- ◯ 3:00am
- ◯ 3:30am
- ◯ 4:00am

25

What time did you wake up TODAY (to the nearest half hour)?

- ◯ 5:00am
- ◯ 5:30am
- ◯ 6:00am
- ◯ 6:30am
- ◯ 7:00am
- ◯ 7:30am
- ◯ 8:00am
- ◯ 8:30am
- ◯ 9:00am

### THE LAST WEEK

NOW think about what you did in the last 7 days...

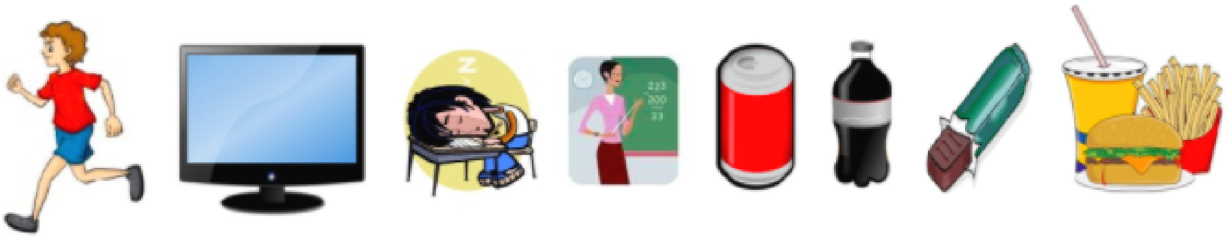

26

In the last 7 days, how many days did you do sports or exercise for at least 1 hour in total (This includes doing any activities or playing sports where your heart beat faster, you breathed faster and you felt warmer?

- ◯ 0 days
- ◯ 1-2 days
- ◯ 3-4 days
- ◯ 5-6 days
- ◯ 7 days

27

In the last 7 days, how many days did you watch TV/play online games/use the internet etc. for 2 or more hours a day (in total)?

- ◯ 0 days
- ◯ 1-2 days
- ◯ 3-4 days
- ◯ 5-6 days
- ◯ 7 days

28

In the last 7 days, how many days did you feel tired?

- ◯ 0 days
- ◯ 1-2 days
- ◯ 3-4 days
- ◯ 5-6 days
- ◯ 7 days

29

In the last 7 days, how many days did you feel like you could concentrate/pay attention well in class?

- ◯ 0 days
- ◯ 1-2 days
- ◯ 3-4 days
- ◯ 5-6 days
- ◯ 7 days

30

In the last 7 days, how many days did you drink at least one fizzy drink (e.g. coke, fanta, sprite)

- 0 days
- 1-2 days
- 3-4 days
- 5-6 days
- 7 days

31

In the last 7 days, how many days did you eat at least one sugary snack (e.g. chocolate bar, sweets)

- 0 days
- 1-2 days
- 3-4 days
- 5-6 days
- 7 days

32

In the last 7 days, how many days did you eat take away foods (e.g. McDonalds, KFC, chinese)

- 0 days
- 1-2 days
- 3-4 days
- 5-6 days
- 7 days

33

In the last 7 days, how many days did your friends or other people you don't live with visit you in your house?

- 0 days
- 1-2 days
- 3-4 days
- 5-6 days
- 7 days

34

In the last 7 days, how many days did you go to your friend's house or someone else's house? (such as someone in your family you don't live with)

- ◯ 0 days
- ◯ 1-2 days
- ◯ 3-4 days
- ◯ 5-6 days
- ◯ 7 days

35

In the last 7 days, have you been ill with a cold?

- ◯ Yes
- ◯ No

### Sport and Activity

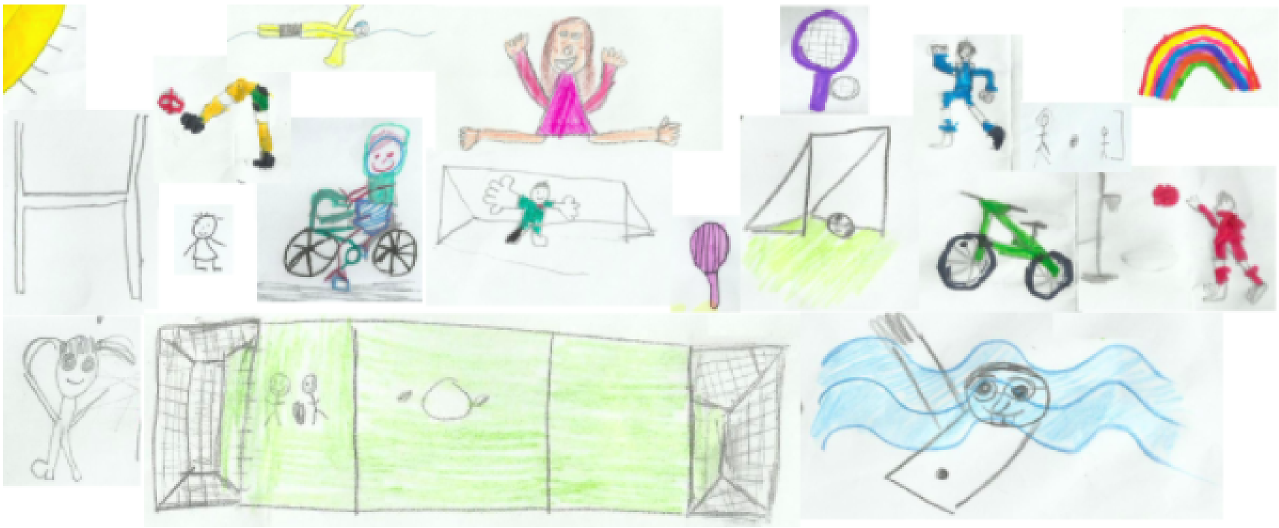

36

These questions are going to ask you how you feel about physical activity (This includes any activity where your heart beats faster, you breathe faster and you feel warmer)

| | Strongly agree | Agree | Disagree | Strongly disagree |
|---|---|---|---|---|
| I want to take part in physical activity | ○ | ○ | ○ | ○ |
| I feel confident to take part in lots of different physical activities | ○ | ○ | ○ | ○ |
| I am good at lots of different physical activities | ○ | ○ | ○ | ○ |
| I understand why taking part in physical activity is good for me | ○ | ○ | ○ | ○ |

37

How many times do you take part in a sports club OUTSIDE OF SCHOOL each week?

- ◯ 0
- ◯ 1
- ◯ 2
- ◯ 3
- ◯ 4
- ◯ 5
- ◯ 6
- ◯ 7
- ◯ 8
- ◯ 9
- ◯ 10

38

Can you ride a bike WITHOUT STABILISERS?

- ◯ Yes
- ◯ No

Can you swim 25 metres WITHOUT A FLOAT OR ARMBANDS? (This is 1 length of a standard swimming pool)

- ◯ Yes
- ◯ No

### You and Your Feelings

This part of the survey is going to ask you how you feel. There are no right or wrong answers. You should just pick the answer which is best for you.

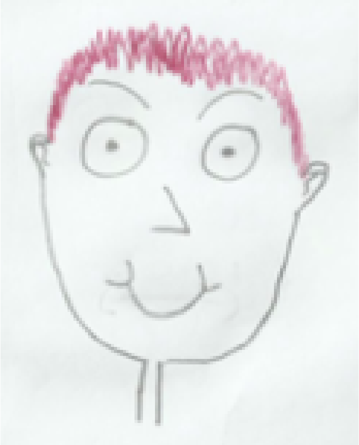
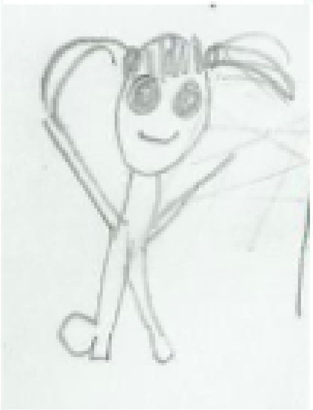
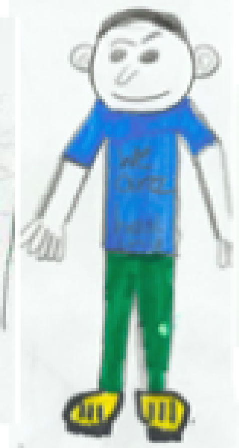
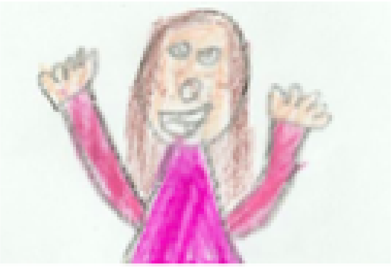

40

#### Tell us if you agree or disagree with the following:

| | Strongly agree | Agree | Don't agree or disagree | Disagree | Strongly disagree |
|---|---|---|---|---|---|
| I am doing well at school | ○ | ○ | ○ | ○ | ○ |
| I feel part of my school community | ○ | ○ | ○ | ○ | ○ |
| I have lots of choice over things that are important to me | ○ | ○ | ○ | ○ | ○ |
| There are lots of things I'm good at | ○ | ○ | ○ | ○ | ○ |

41

On a scale of 1 to 10 (1 being very unhappy and 10 being very happy, how do you feel about:

Your Health

☆ ☆ ☆ ☆ ☆ ☆ ☆ ☆ ☆ ☆

42

On a scale of 1 to 10 (1 being very unhappy and 10 being very happy, how do you feel about:

Your School

☆ ☆ ☆ ☆ ☆ ☆ ☆ ☆ ☆ ☆

43

On a scale of 1 to 10 (1 being very unhappy and 10 being very happy, how do you feel about:

Your Family

☆ ☆ ☆ ☆ ☆ ☆ ☆ ☆ ☆ ☆

44

On a scale of 1 to 10 (1 being very unhappy and 10 being very happy, how do you feel about:

Your Friends

☆ ☆ ☆ ☆ ☆ ☆ ☆ ☆ ☆ ☆

45

On a scale of 1 to 10 (1 being very unhappy and 10 being very happy, how do you feel about:

Your Appearance (how you look)

☆ ☆ ☆ ☆ ☆ ☆ ☆ ☆ ☆ ☆

46

On a scale of 1 to 10 (1 being very unhappy and 10 being very happy, how do you feel about:

Your Life

☆ ☆ ☆ ☆ ☆ ☆ ☆ ☆ ☆ ☆

### You and Your Feelings

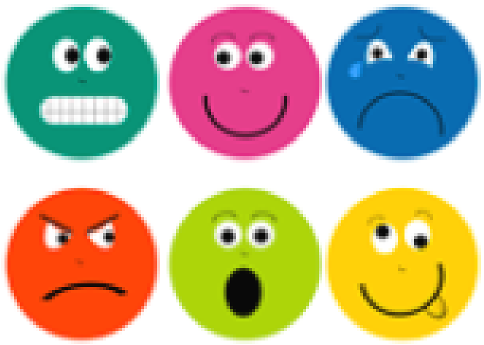

Based on the Me and My Feelings Questionnaire ( Deighton, Tymms, Vostanis, Belsky, Fonagy, Brown, Martin, Patalay, & Wolpert, 2012)

Remember, there are no right or wrong answers, just pick which is right for you.

| | Never | Sometimes | Always |
|---|---|---|---|
| I feel lonely | ◯ | ◯ | ◯ |
| I cry a lot | ◯ | ◯ | ◯ |
| I am unhappy | ◯ | ◯ | ◯ |
| I feel nobody likes me | ◯ | ◯ | ◯ |
| I worry a lot | ◯ | ◯ | ◯ |
| I have problems sleeping | ◯ | ◯ | ◯ |
| I wake up in the night | ◯ | ◯ | ◯ |
| I am shy | ◯ | ◯ | ◯ |
| I feel scared | ◯ | ◯ | ◯ |
| I worry when I am at school | ◯ | ◯ | ◯ |
| I get very angry | ◯ | ◯ | ◯ |
| I lose my temper | ◯ | ◯ | ◯ |
| I hit out when I am angry | ◯ | ◯ | ◯ |
| I do things to hurt people | ◯ | ◯ | ◯ |
| I am calm | ◯ | ◯ | ◯ |
| I break things on purpose | ◯ | ◯ | ◯ |

### Your Local Area

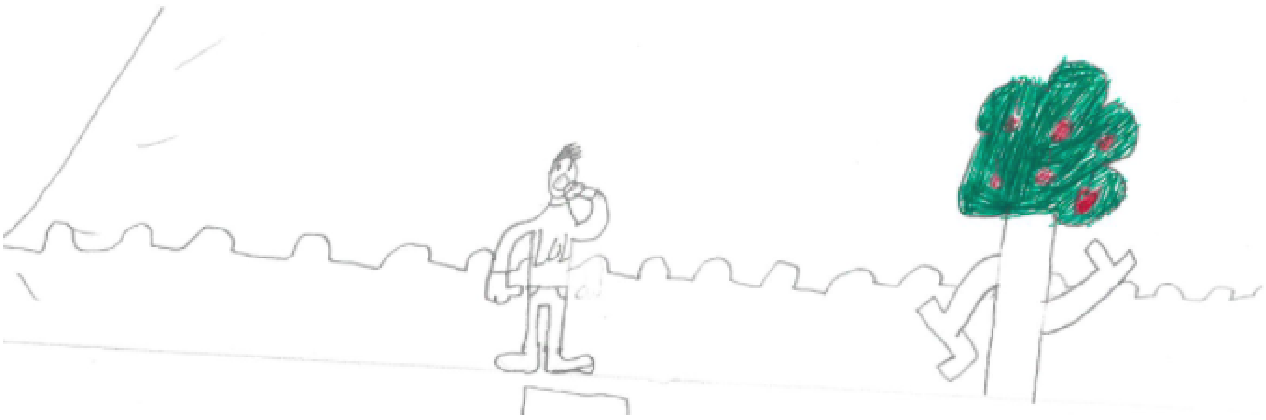

48

On a scale of 1 to 10 (1 being not very safe and 10 being very safe), how safe do you feel playing in your area?

☆ ☆ ☆ ☆ ☆ ☆ ☆ ☆ ☆ ☆

49

From your house, can you walk to school?

◯ Yes

◯ No

50

From your house, can you easily walk to a park?

◯ Yes

◯ No

51

From your house, can you easily walk to a leisure centre/sports centre?

- Yes
- No

52

Can you play in all the places you would like to?

- I can play in all the places I would like to
- I can play in some of the places I would like to
- I can only play in a few places I would like to
- I can hardly play in any of the places I would like to

53

Are you happy with the area that you live in?

- Yes
- No

54

If you could change something to make you and your friends healthier and happier, what would you change... IN SCHOOL?

55

If you could change something to make you and your friends healthier and happier, what would you change... OUT OF SCHOOL?

### Don't forget to press submit below!

We have some resources on our website if you would like to learn more or would like to speak to someone... https://happen-wales.co.uk/some-resources-for-you/ (https://happen-wales.co.uk/some-resources-for-you/)
